# Effect of Dexamethasone in Hospitalized Patients with COVID-19 – Preliminary Report

**DOI:** 10.1101/2020.06.22.20137273

**Authors:** Peter Horby, Wei Shen Lim, Jonathan Emberson, Marion Mafham, Jennifer Bell, Louise Linsell, Natalie Staplin, Christopher Brightling, Andrew Ustianowski, Einas Elmahi, Benjamin Prudon, Christopher Green, Timothy Felton, David Chadwick, Kanchan Rege, Christopher Fegan, Lucy C Chappell, Saul N Faust, Thomas Jaki, Katie Jeffery, Alan Montgomery, Kathryn Rowan, Edmund Juszczak, J Kenneth Baillie, Richard Haynes, Martin J Landray, RECOVERY Collaborative Group

**Author notes:** The writing committee and trial steering committee are listed at the end of this manuscript and a complete list of collaborators in the Randomised Evaluation of COVID-19 Therapy (RECOVERY) trial is provided in the Supplementary Appendix. Correspondence to: Dr Peter W Horby and Dr Martin J Landray, RECOVERY Central Coordinating Office, Richard Doll Building, Old Road Campus, Roosevelt Drive, Oxford OX3 7LF, United Kingdom.

## Abstract

**Background:** Coronavirus disease 2019 (COVID-19) is associated with diffuse lung damage. Corticosteroids may modulate immune-mediated lung injury and reducing progression to respiratory failure and death.

**Methods:** The Randomised Evaluation of COVID-19 therapy (RECOVERY) trial is a randomized, controlled, open-label, adaptive, platform trial comparing a range of possible treatments with usual care in patients hospitalized with COVID-19. We report the preliminary results for the comparison of dexamethasone 6 mg given once daily for up to ten days vs. usual care alone. The primary outcome was 28-day mortality.

**Results:** 2104 patients randomly allocated to receive dexamethasone were compared with 4321 patients concurrently allocated to usual care. Overall, 454 (21.6%) patients allocated dexamethasone and 1065 (24.6%) patients allocated usual care died within 28 days (age-adjusted rate ratio [RR] 0.83; 95% confidence interval [CI] 0.74 to 0.92; P<0.001). The proportional and absolute mortality rate reductions varied significantly depending on level of respiratory support at randomization (test for trend p<0.001): Dexamethasone reduced deaths by one-third in patients receiving invasive mechanical ventilation (29.0% vs. 40.7%, RR 0.65 [95% CI 0.51 to 0.82]; p<0.001), by one-fifth in patients receiving oxygen without invasive mechanical ventilation (21.5% vs. 25.0%, RR 0.80 [95% CI 0.70 to 0.92]; p=0.002), but did not reduce mortality in patients not receiving respiratory support at randomization (17.0% vs. 13.2%, RR 1.22 [95% CI 0.93 to 1.61]; p=0.14).

**Conclusions:** In patients hospitalized with COVID-19, dexamethasone reduced 28-day mortality among those receiving invasive mechanical ventilation or oxygen at randomization, but not among patients not receiving respiratory support.

**Trial registrations:** The RECOVERY trial is registered with ISRCTN (50189673) and clinicaltrials.gov (NCT04381936).

**Funding:** Medical Research Council and National Institute for Health Research (Grant ref: MC_PC_19056).

## INTRODUCTION

Severe acute respiratory syndrome coronavirus 2 (SARS-CoV-2), the cause of coronavirus disease 2019 (COVID-19), emerged in China in late 2019 from a zoonotic source.^1^ The majority of COVID-19 infections are either asymptomatic or result in only mild disease. However, a substantial proportion of older infected individuals develop a respiratory illness requiring hospital care,^2^ which can progress to critical illness with hypoxemic respiratory failure requiring prolonged ventilatory support.^3-6^ Amongst COVID-19 patients admitted to UK hospitals, the case fatality rate is over 26%, and is over 37% in patients requiring invasive mechanical ventilation.^7^ Although remdesivir has been shown to shorten the time to recovery in hospitalized patients,^8^ no therapeutic agents have been shown to reduce mortality.

The pathophysiology of severe COVID-19 is dominated by an acute pneumonic process with extensive radiological opacity and, on autopsy, diffuse alveolar damage, inflammatory infiltrates and microvascular thromobosis.^9,10^ The host immune response is thought to play a key role in the pathophysiology of organ failure in other severe viral pneumonias such as highly pathogenic avian influenza,^11^ severe acute respiratory syndrome (SARS),^12,13^ and pandemic and seasonal influenza.^14^ Inflammatory organ injury may occur in severe COVID-19, with a subset of patients having markedly elevated inflammatory markers such as C-reactive protein, ferritin, and interleukins 1 and 6.^6,15,16^ Several therapeutic interventions to mitigate inflammatory organ injury have been proposed in viral pneumonia but the value of corticosteroids has been widely debated.^17,18^

In the absence of reliable evidence from large-scale randomized clinical trials, there is great uncertainty about the effectiveness of corticosteroids in COVID-19. Prior to RECOVERY, many COVID-19 treatment guidelines stated that corticosteroids were either ‘contraindicated’ or ‘not recommended’^19^ although in China, corticosteroids are recommended for severe cases.^20^ Practice has varied widely across the world: in some series, as many as 50% of cases were treated with corticosteroids.^21,22^ Here we report the results of a randomized controlled trial of dexamethasone in patients hospitalized with COVID-19.

## METHODS

### Trial design and participants

The RECOVERY trial is an investigator-initiated, individually randomized, controlled, open-label, adaptive platform trial to evaluate the effects of potential treatments in patients hospitalized with COVID-19. The trial was conducted at 176 National Health Service (NHS) hospital organizations in the United Kingdom (see Supplementary Appendix), supported by the National Institute for Health Research Clinical Research Network. The trial was coordinated by the Nuffield Department of Population Health at University of Oxford, the trial sponsor.

Hospitalized patients were eligible for the trial if they had clinically suspected or laboratory confirmed SARS-CoV-2 infection and no medical history that might, in the opinion of the attending clinician, put the patient at significant risk if they were to participate in the trial. Initially, recruitment was limited to patients aged at least 18 years but the age limit was removed from 9 May 2020. Pregnant or breast-feeding women were eligible.

Written informed consent was obtained from all patients or from a legal representative if they were too unwell or unable to provide consent. The trial was conducted in accordance with the principles of the International Conference on Harmonization–Good Clinical Practice guidelines and approved by the UK Medicines and Healthcare Products Regulatory Agency (MHRA) and the Cambridge East Research Ethics Committee (ref: 20/EE/0101). The protocol and statistical analysis plan are available on the study website www.recoverytrial.net.

### Randomization

Baseline data collected using a web-based case report form included demographics, level of respiratory support, major comorbidities, suitability of the study treatment for a particular patient and treatment availability at the study site. Eligible and consenting patients were assigned in a ratio of 2:1 to either usual standard of care or to usual standard of care plus dexamethasone 6 mg once daily (oral or intravenous) for up to 10 days (or until discharge if sooner) or to one of the other suitable and available treatment arms (see Supplementary Appendix) using web-based simple randomization with allocation concealment. For some patients, dexamethasone was either unavailable at the hospital at the time of enrolment or considered by the managing doctor to be either definitely indicated or definitely contraindicated. These patients were excluded from entry in the randomized comparison of dexamethasone vs. usual care and hence are not part of this report. The randomly assigned treatment was prescribed by the treating clinician. Participants and local study staff were not blinded to the allocated treatment.

### Procedures

A single online follow-up form was to be completed when participants were discharged, had died or at 28 days after randomization (whichever occurred earlier). Information was recorded on adherence to allocated study treatment, receipt of other study treatments, duration of admission, receipt of respiratory or renal support, and vital status (including cause of death). In addition, routine health care and registry data were obtained including information on vital status (with date and cause of death); discharge from hospital; intensive care use; and renal replacement therapy.

### Outcome measures

The primary outcome was all-cause mortality within 28 days of randomization. Secondary outcomes were time to discharge from hospital, and among patients not receiving invasive mechanical ventilation at randomization, subsequent receipt of invasive mechanical ventilation (including extra-corporeal membrane oxygenation) or death. Subsidiary clinical outcomes included cause-specific mortality, receipt of renal hemodialysis or hemofiltration, major cardiac arrhythmia (recorded in a subset), and receipt and duration of ventilation.

### Statistical Analysis

For the primary outcome of 28-day mortality, the hazard ratio from Cox regression was used to estimate the mortality rate ratio. The few patients (4.8%) who had not been followed for 28 days by the time of the data cut (10 June 2020) were either censored on 8 June 2020 or, if they had already been discharged alive, were right-censored at day 29 (that is, in the absence of any information to the contrary they were assumed to have survived 28 days). Kaplan-Meier survival curves were constructed to display cumulative mortality over the 28-day period. Cox regression was used to analyze the secondary outcome of hospital discharge within 28 days, with patients who died in hospital right-censored on day 29. For the pre-specified composite secondary outcome of invasive mechanical ventilation or death within 28 days (among those not receiving invasive mechanical ventilation at randomization), the precise date of invasive mechanical ventilation was not available and so a log-binomial regression model was used to estimate the risk ratio.

Pre-specified analyses of the primary outcome were performed in five subgroups defined by characteristics at randomization: age, sex, level of respiratory support, days since symptom onset, and predicted 28-day mortality risk. Observed effects within subgroup categories were compared using a chi-square test for trend. Through the play of chance in the unstratified randomization, mean age was 1.1 years higher in those allocated dexamethasone than those allocated usual care (Table 1). To account for this imbalance in an important prognostic factor, the estimates of rate ratios and risk ratios (both hereon denoted RR) were adjusted for baseline age. This adjustment was not specified in the first version of the statistical analysis plan, but was added once the imbalance in age (a key prognostic factor) became apparent. Results with and without age-adjustment are provided and show that it does not alter the conclusions materially.

**Table 1:** Baseline characteristics by randomized allocation and level of respiratory support received

|  | Treatment allocation |  | Respiratory support received at randomization |  |  |
| --- | --- | --- | --- | --- | --- |
|  | Dexamethasone (n=2104) | Usual care (n=4321) | No oxygen received (n=1535) | Oxygen only (n=3883) | Invasive mechanical ventilation (n=1007) |
| Age, years | 66.9 (15.4) | 65.8 (15.8) | 69.3 (17.6) | 66.7 (15.3) | 59.0 (11.5) |
| <70 | 1142 (54%) | 2506 (58%) | 660 (43%) | 2149 (55%) | 839 (83%) |
| ≥70 to <80 | 467 (22%) | 860 (20%) | 338 (22%) | 837 (22%) | 152 (15%) |
| ≥80 | 495 (24%) | 955 (22%) | 537 (35%) | 897 (23%) | 16 (2%) |
| Sex |  |  |  |  |  |
| Male | 1338 (64%) | 2750 (64%) | 892 (58%) | 2462 (63%) | 734 (73%) |
| Female* | 766 (36%) | 1571 (36%) | 643 (42%) | 1421 (37%) | 273 (27%) |
| Number of days since symptom onset | 8 (5-13) | 9 (5-13) | 6 (3-10) | 9 (5-12) | 13 (8-18) |
| Respiratory support received |  |  |  |  |  |
| No oxygen received | 501 (24%) | 1034 (24%) | 1535 (100%) | 0 (0%) | 0 (0%) |
| Oxygen only | 1279 (61%) | 2604 (60%) | 0 (0%) | 3883 (100%) | 0 (0%) |
| Invasive mechanical ventilation | 324 (15%) | 683 (16%) | 0 (0%) | 0 (0%) | 1007 (100%) |
| Previous diseases |  |  |  |  |  |
| Diabetes | 521 (25%) | 1025 (24%) | 342 (22%) | 950 (24%) | 254 (25%) |
| Heart disease | 586 (28%) | 1171 (27%) | 519 (34%) | 1074 (28%) | 164 (16%) |
| Chronic lung disease | 415 (20%) | 931 (22%) | 351 (23%) | 883 (23%) | 112 (11%) |
| Tuberculosis | 6 (<0.5%) | 19 (<0.5%) | 8 (1%) | 11 (<0.5%) | 6 (1%) |
| HIV | 12 (1%) | 20 (<0.5%) | 5 (<0.5%) | 21 (1%) | 6 (1%) |
| Severe liver disease | 37 (2%) | 82 (2%) | 32 (2%) | 72 (2%) | 15 (1%) |
| Severe kidney impairment | 167 (8%) | 358 (8%) | 120 (8%) | 253 (7%) | 152 (15%) |
| Any of the above | 1174 (56%) | 2417 (56%) | 911 (59%) | 2175 (56%) | 505 (50%) |
| SARS-Cov-2 test result |  |  |  |  |  |
| Positive | 1702 (81%) | 3553 (82%) | 1198 (78%) | 3144 (81%) | 913 (91%) |
| Negative | 213 (10%) | 397 (9%) | 182 (12%) | 398 (10%) | 30 (3%) |
| Test result not yet known† | 189 (9%) | 371 (9%) | 155 (10%) | 341 (9%) | 64 (6%) |

Results are count (%), mean ± standard deviation, or median (inter-quartile range). \* Includes 6 pregnant women. † SARS-Cov-2 test results are captured on the follow-up form, so are currently unknown for some. There was a significant (2p=0.008) difference in mean age between those allocated dexamethasone and those allocated usual care, but no significant differences between these groups in any other baseline characteristic. The 'oxygen only' group includes non-invasive ventilation.

Estimates of rate and risk ratios are shown with 95% confidence intervals. All p-values are 2-sided and all analyses were done according to the intention-to-treat principle. The full database is held by the study team which collected the data from study sites and performed the analyses at the Nuffield Department of Population Health, University of Oxford.

### Sample size and decision to stop enrolment

As stated in the protocol, appropriate sample sizes could not be estimated when the trial was being planned at the start of the COVID-19 pandemic. As the trial progressed, the trial Steering Committee, blinded to the results of the study treatment comparisons, formed the view that, if 28-day mortality was 20% then a comparison of at least 2000 patients allocated to active drug and 4000 to usual care alone would yield at least 90% power at two-sided P=0.01 to detect a clinically relevant absolute difference of 4 percentage points between the two groups (a proportional reduction of one-fifth). Consequently, on 8 June 2020, the Steering Committee closed recruitment to the dexamethasone arm since enrolment exceeded 2000 patients.

## RESULTS

### Patients

Of the 11,320 patients randomized between 19 March and 8 June, 9355 (83%) were eligible to be randomized to dexamethasone (that is dexamethasone was available in the hospital at the time and the patient had no known indication for or contraindication to dexamethasone). Of these, 2104 were randomized to dexamethasone and 4321 were randomized to usual care (Figure S1), with the remainder being randomized to one of the other treatment arms. Mean age of study participants in this comparison was 66.1 years (Table 1) and 36% patients were female. A history of diabetes was present in 24% of patients, heart disease in 27%, and chronic lung disease in 21%, with 56% having at least one major comorbidity recorded. In this analysis, 82% of patients had laboratory confirmed SARS-CoV-2 infection, with the result currently awaited for 9%. At randomization, 16% were receiving invasive mechanical ventilation or extracorporeal membrane oxygenation, 60% were receiving oxygen only (with or without non-invasive ventilation), and 24% were receiving neither.

Follow-up information was complete for 6119 (95%) of the randomized patients. Of those allocated to dexamethasone 95% received at least 1 dose (Table S1) and the median number of days of treatment was 6 days. 7% of the usual care group received dexamethasone. Use of azithromycin during the follow-up period was similar in both arms (23% vs. 24%) and very few patients received hydroxychloroquine, lopinavir-ritonavir, or interleukin-6 antagonists during follow-up (Table S1). Remdesivir only became available for use in the UK under the MHRA Emergency Access to Medicines Scheme on 26 May 2020.

### Primary outcome

Significantly fewer patients allocated to dexamethasone met the primary outcome of 28-day mortality than in the usual care group (454 of 2104 patients [21.6%] allocated dexamethasone vs. 1065 of 4321 patients [24.6%] allocated usual care; rate ratio, 0.83; 95% confidence interval [CI], 0.74 to 0.92; P<0.001) (Figure 1A). In a pre-specified subgroup analysis by level of respiratory support received at randomization, there was a significant trend showing the greatest absolute and proportional benefit among those patients receiving invasive mechanical ventilation at randomization (test for trend p<0.001) (Figure 2). Dexamethasone reduced 28-day mortality by 35% in patients receiving invasive mechanical ventilation (rate ratio 0.65 [95% CI 0.51 to 0.82]; p<0.001) and by 20% in patients receiving oxygen without invasive mechanical ventilation (rate ratio 0.80 [95% CI 0.70 to 0.92]; p=0.002) (Figure 1B-C). However, there was no evidence of benefit among those patients who were not receiving respiratory support (rate ratio 1.22 [95% CI 0.93 to 1.61]; p=0.14) (Figure 1D). Sensitivity analyses without age-adjustment produced similar findings (Table S2).

**Figure 1:**
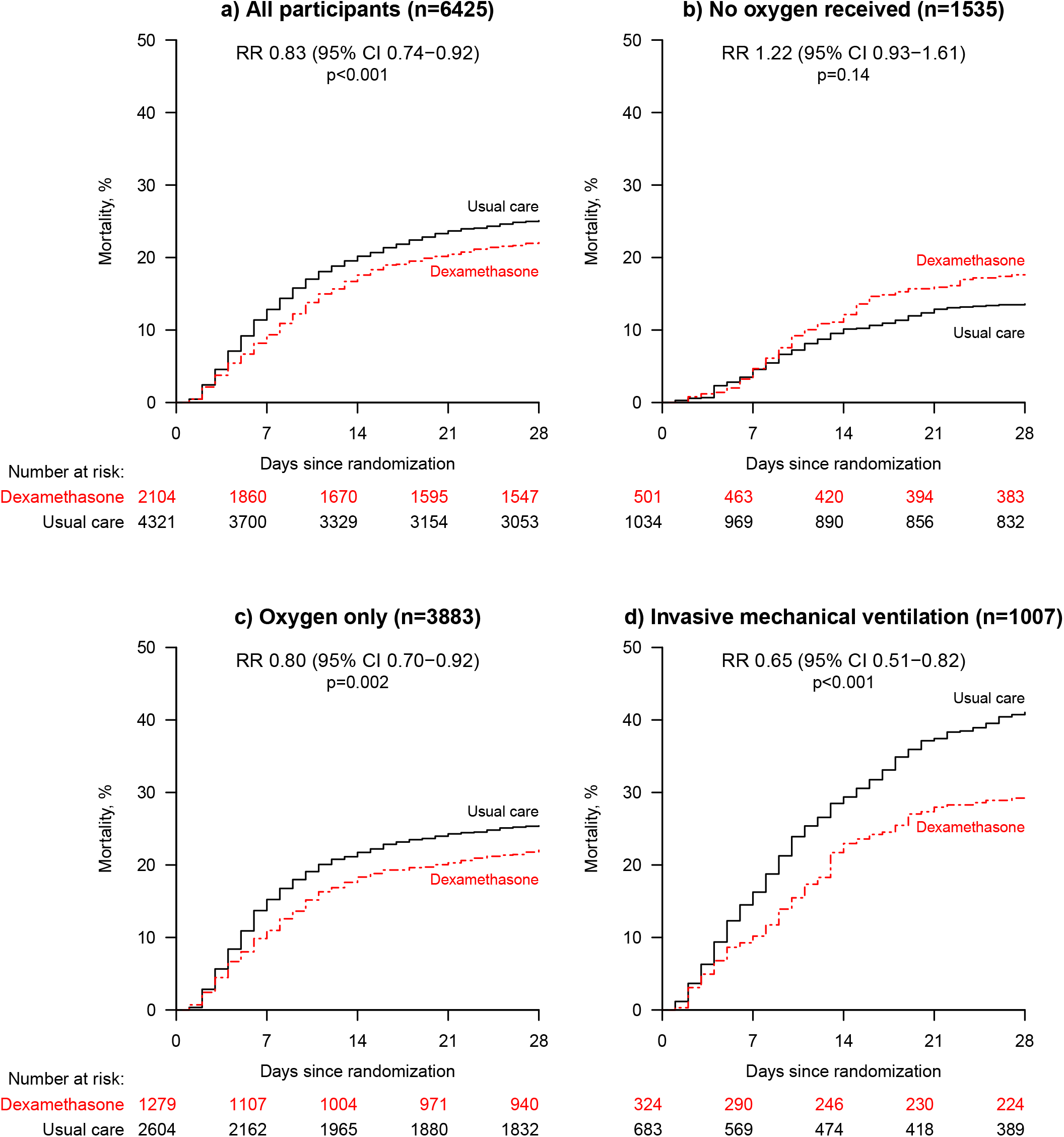
28-day mortality in all patients (panel a) and separately according to level of respiratory support received at randomization (panels b-d) RR=age-adjusted rate ratio. CI=confidence interval. The ‘oxygen only’ group includes non-invasive ventilation. Note: in the RECOVERY trial press release of 16 June 2020, effects in subgroups of level of respiratory support received were shown with 99% CIs, not 95% CIs as inadvertently stated. The age-adjusted rate ratio and 99% confidence intervals remain unchanged in this analysis: no oxygen required, RR 1.22 (99% CI 0.86-1.75); oxygen only, RR 0.80 (99% CI 0.67-0.96); invasive mechanical ventilation, RR 0.65 (99% CI 0.48-0.88).

**Figure 2:**
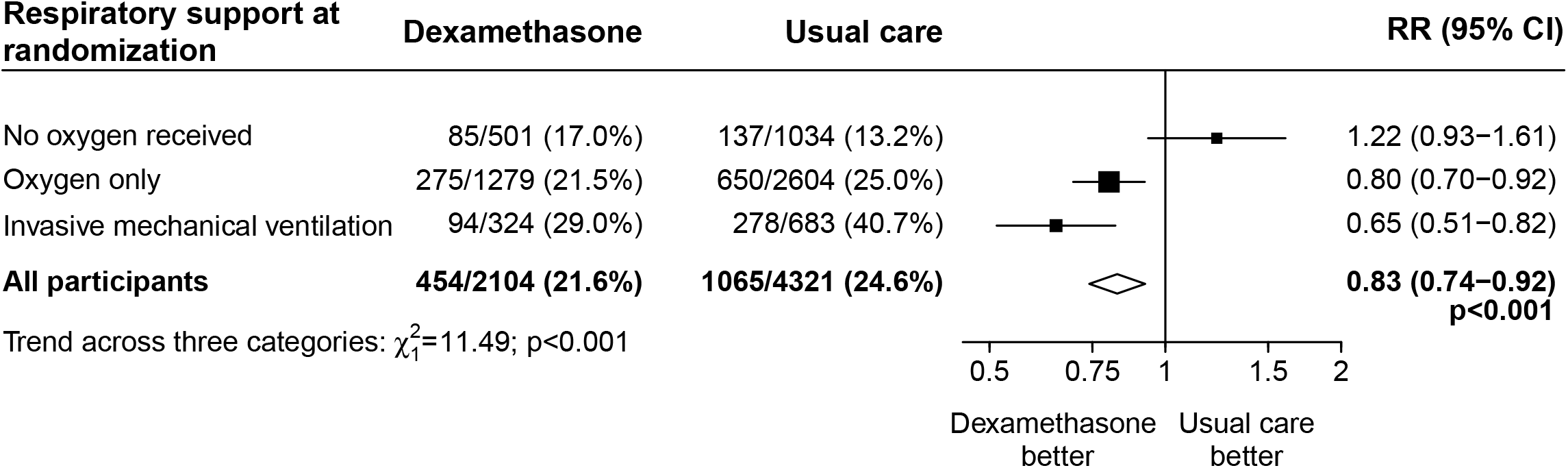
Effect of allocation to dexamethasone on 28-day mortality by level of respiratory support received at randomization. RR=age-adjusted rate ratio. CI=confidence interval. Subgroup-specific RR estimates are represented by squares (with areas of the squares proportional to the amount of statistical information) and the lines through them correspond to the 95% confidence intervals. The ‘oxygen only’ group includes non-invasive ventilation. Note: in the RECOVERY trial press release of 16 June 2020, effects in subgroups of level of respiratory support received were shown with 99% CIs, not 95% CIs as inadvertently stated. The age-adjusted rate ratio and 99% confidence intervals remain unchanged in this analysis: no oxygen required, RR 1.22 (99% CI 0.86-1.75); oxygen only, RR 0.80 (99% CI 0.67-0.96); invasive mechanical ventilation, RR 0.65 (99% CI 0.48-0.88).

Patients on invasive mechanical ventilation at randomization were on average 10 years younger than those not receiving any respiratory support and had symptoms prior to randomization for 7 days longer (Table 1). 28-day mortality in the usual care group was highest in those who were receiving invasive mechanical ventilation at randomization (40.7%), intermediate in those patients who received oxygen only (25.0%), and lowest among those who were not receiving respiratory support at randomization (13.2%). Consequently, the greatest absolute reductions in 28-day mortality were seen among those patients on invasive mechanical ventilation.

Patients with longer duration of symptoms (who were more likely to be on invasive mechanical ventilation at randomization) had a greater mortality benefit, such that dexamethasone was associated with a reduction in 28-day mortality among those with symptoms for more than 7 days but not among those with more recent symptom onset (test for trend p<0.001) (Figure S2).

### Secondary outcomes

Allocation to dexamethasone was associated with a shorter duration of hospitalization than usual care (median 12 days vs. 13 days) and a greater probability of discharge within 28 days (rate ratio 1.11 [95% CI 1.04 to 1.19]; p=0.002) (Table 2) with the greatest effect seen among those receiving invasive mechanical ventilation at baseline (test for trend p=0.002) (Figure S3a).

**Table 2:** Effect of allocation to dexamethasone on main study outcomes

|  | Treatment allocation |  | RR (95% CI) | p-value |
| --- | --- | --- | --- | --- |
|  | Dexamethasone<br>(n=2104) | Usual care<br>(n=4321) |  |  |
| Primary outcome: |  |  |  |  |
| 28-day mortality | 454 (21.6%) | 1065 (24.6%) | 0.83 (0.74-0.92) | <0.001 |
| Secondary outcomes: |  |  |  |  |
| Discharged from hospital within 28 days | 1360 (64.6%) | 2639 (61.1%) | 1.11 (1.04-1.19) | 0.002 |
| Receipt of invasive mechanical ventilation or death* | 425/1780 (23.9%) | 939/3638 (25.8%) | 0.91 (0.82-1.00) | 0.049 |
| Invasive mechanical ventilation | 92/1780 (5.2%) | 258/3638 (7.1%) | 0.76 (0.61-0.96) | 0.021 |
| Death | 360/1780 (20.2%) | 787/3638 (21.6%) | 0.91 (0.82-1.01) | 0.07 |

RR=Rate Ratio for the outcomes of 28-day mortality and hospital discharge, and risk ratio for the outcome of receipt of invasive mechanical ventilation or death (and its subcomponents). Estimates of the RR and its 95% confidence interval are adjusted for age in three categories (<70 years , 70-79 years, and 80 years or older). \* Analyses exclude those on invasive mechanical ventilation at randomization.

Among those not on invasive mechanical ventilation at baseline, the number of patients progressing to the pre-specified composite secondary outcome of invasive mechanical ventilation or death was lower among those allocated to dexamethasone (risk ratio 0.91 [95% CI 0.82 to 1.00]; p=0.049) (Table 2) but with significantly greater effects among patients receiving oxygen at randomization (test for trend p=0.008) (Figure S3b).

### Subsidiary clinical outcomes

The risk of progression to invasive mechanical ventilation was lower among those allocated dexamethasone vs. usual care (risk ratio 0.76 [95% CI 0.61 to 0.96]; p=0.021) (Table 2). Preliminary analyses indicate no excess risk of any particular cause of death (in particular there was no excess of deaths due to non-COVID infection). More detailed analyses of cause-specific mortality, need for renal dialysis or hemofiltration, and duration of ventilation are in preparation.

## DISCUSSION

These preliminary results show that dexamethasone 6mg per day for up to 10 days reduces 28-day mortality in COVID-19 patients receiving invasive mechanical ventilation by one third, and by one fifth in patients receiving oxygen without invasive mechanical ventilation. Similarly, benefit was clearer in patients treated more than 7 days after treatment onset, when inflammatory lung damage is likely to have been more common. However, no benefit was demonstrated in hospitalized COVID-19 patients who were not receiving respiratory support and the results are consistent with possible harm in this group.

RECOVERY is a large, pragmatic, randomized, controlled adaptive platform trial designed to provide rapid and robust assessment of the impact of readily available potential treatments for COVID-19 on 28-day mortality. Around 15% of all UK hospitalized patients with COVID-19 were enrolled in the trial and the control arm fatality rate is consistent with the overall hospitalized case fatality rate in the UK.^7^ Only essential data were collected at hospital sites with additional information (including long-term mortality) ascertained through linkage with routine data sources. We did not collect information on physiological, laboratory or virologic parameters. The protocol combines the methods of large, simple trials of treatments for acute myocardial infarction in the 1980s with the opportunities provided by digital health care in the 2020s.^23-25^ It has progressed at unprecedented speed, as is essential for studies during epidemics.^26^ These preliminary results for dexamethasone were announced on 16 June 2020, just 98 days after the protocol was first drafted, and were adopted into UK practice later the same day.^27^

Corticosteroids have been widely used in syndromes closely related to COVID-19, including SARS, MERS, severe influenza, and community acquired pneumonia. However, the evidence to support or discourage the use of corticosteroids in these conditions has been very weak due to the lack of sufficiently powered randomized controlled trials.^28-31^ In addition, the evidence base has suffered from heterogeneity in corticosteroid doses, medical conditions, and disease severity studied. It is likely that the beneficial effect of corticosteroids in severe viral respiratory infections is dependent on using the right dose, at the right time, in the right patient. High doses may be more harmful than helpful, as may corticosteroid treatment given at a time when control of viral replication is paramount and inflammation is minimal. Slower clearance of viral RNA has been observed in patients with SARS, MERS and influenza treated with systemic corticosteroids but the clinical significance of this is unknown.^29,32,33^ Unlike SARS, where viral replication peaks in the second week of illness,^34^ peak viral shedding in COVID-19 appears to be higher early in the illness and declines thereafter.^35-38^ The greater mortality benefit of dexamethasone in patients with COVID-19 who required respiratory support, and among those recruited after the first week of their illness, suggests that at this stage the disease is dominated by immunopathology, with active virus replication playing a secondary role. It is also possible there is an effect via mineralocorticoid receptor binding in the context of SARS-CoV-2 induced dysregulation of the renin-angiotensin system.^39^ This would caution against extrapolating the effect of dexamethasone in patients with COVID-19 to patients with other viral respiratory diseases that have a different natural history.

The RECOVERY trial provides clear evidence that treatment with dexamethasone 6 mg once daily for up to 10 days reduces 28-day mortality in patients with COVID-19 who are receiving respiratory support. Based on these results, 1 death would be prevented by treatment of around 8 patients requiring invasive mechanical ventilation or around 25 patients requiring oxygen (which, in the UK, is recommended when oxygen saturations on room air are 92-94%)^40^ without invasive mechanical ventilation. There was no benefit (and the possibility of harm) among patients who did not require oxygen. Prior to the completion of this trial, many COVID-19 treatment guidelines have stated that corticosteroids are either ‘contraindicated’ or ‘not recommended’ in COVID-19.^19^ These should now be updated, as has already happened within the UK.^27^ Dexamethasone provides an effective treatment for the sickest patients with COVID-19 and, given its low cost, well understood safety profile, and widespread availability, is one that can be used worldwide.

## Data Availability

The protocol, consent form, statistical analysis plan, definition & derivation of clinical characteristics & outcomes, training materials, regulatory documents, and other relevant study materials are available online at www.recoverytrial.net
This is a preliminary report and follow-up is ongoing. Data will be made available to bona fide researchers registered with an appropriate institution within 3 months of the final participant completing 28-day follow-up.
As described in the protocol, the trial Steering Committee will facilitate the use of the study data and approval will not be unreasonably withheld. However, the Steering Committee will need to be satisfied that any proposed publication is of high quality, honours the commitments made to the study participants in the consent documentation and ethical approvals, and is compliant with relevant legal and regulatory requirements (e.g. relating to data protection and privacy). The Steering Committee will have the right to review and comment on any draft manuscripts prior to publication.

## Authorship

This manuscript was prepared by the Writing Committee and reviewed and approved by all members of the trial Steering Committee. The funders had no role in the analysis of the data, preparation and approval of this manuscript, or the decision to submit it for publication. The first and last members of the Writing Committee vouch for the data and analyses, and for the fidelity of this report to the study protocol and data analysis plan.

### Writing Committee (on behalf of the RECOVERY Collaborative Group)

Peter Horby FRCP,^a,^* Wei Shen Lim FRCP,^b,^* Jonathan Emberson PhD,^c,d^ Marion Mafham MD,^c^ Jennifer Bell MSc,^c^ Louise Linsell DPhil,^c^ Natalie Staplin PhD,^c,d^ Christopher Brightling FMedSci,^e^ Andrew Ustianowski PhD,^f^ Einas Elmahi MPhil,^g^ Benjamin Prudon FRCP,^h^ Christopher Green DPhil,^i^ Timothy Felton PhD,^j^ David Chadwick PhD,^k^ Kanchan Rege FRCPath,^l^ Christopher Fegan MD,^m^ Lucy C Chappell PhD,^n^ Saul N Faust FRCPCH,° Thomas Jaki PhD,^p,q^ Katie Jeffery PhD,^r^ Alan Montgomery PhD,^s^ Kathryn Rowan PhD,^t^ Edmund Juszczak PhD,^c^ J Kenneth Baillie MD PhD,^u^ Richard Haynes DM,^c,d†^ Martin J Landray PhD.^c,d,v†^

^a^ Nuffield Department of Medicine, University of Oxford, Oxford, United Kingdom.

^b^ Respiratory Medicine Department, Nottingham University Hospitals NHS Trust, Nottingham, United Kingdom

^c^ Nuffield Department of Population Health, University of Oxford, Oxford, United Kingdom

^d^ MRC Population Health Research Unit, University of Oxford, Oxford, United Kingdom

^e^ Institute for Lung Health, Leicester NIHR Biomedical Research Centre, University of Leicester, Leicester, United Kingdom

^f^ Regional Infectious Diseases Unit, North Manchester General Hospital & University of Manchester, Manchester, United Kingdom

^g^ Research and Development Department, Northampton General Hospital, Northampton, United Kingdom

^h^ Department of Respiratory Medicine, North Tees & Hartlepool NHS Foundation Trust, Stockton-on-Tees, United Kingdom

^i^ University Hospitals Birmingham NHS Foundation Trust and Institute of Microbiology & Infection, University of Birmingham, Birmingham, United Kingdom

^j^ University of Manchester and Manchester University NHS Foundation Trust, Manchester, United Kingdom

^k^ Centre for Clinical Infection, James Cook University Hospital, Middlesbrough, United Kingdom

^l^ North West Anglia NHS Foundation Trust, Peterborough, United Kingdom

^m^ Department of Research and Development, Cardiff and Vale University Health Board, Cardiff, United Kingdom

^n^ School of Life Course Sciences, King’s College London, London, United Kingdom

° NIHR Southampton Clinical Research Facility and Biomedical Research Centre, University Hospital Southampton NHS Foundation Trust and University of Southampton, Southampton, United Kingdom

^p^ Department of Mathematics and Statistics, Lancaster University, Lancaster, United Kingdom

^q^ MRC Biostatistics Unit, University of Cambridge, Cambridge, United Kingdom

^r^ Oxford University Hospitals NHS Foundation Trust, Oxford, United Kingdom

^s^ School of Medicine, University of Nottingham, Nottingham, United Kingdom

^t^ Intensive Care National Audit & Research Centre, London, United Kingdom

^u^ Roslin Institute, University of Edinburgh, Edinburgh, United Kingdom

^v^ NIHR Oxford Biomedical Research Centre, Oxford University Hospitals NHS Foundation Trust, Oxford, United Kingdom

*,^†^ equal contribution

## Data Monitoring Committee

Peter Sandercock, Janet Darbyshire, David DeMets, Robert Fowler, David Lalloo, Ian Roberts, Janet Wittes.

## Acknowledgements

We would like to thank the many thousands of doctors, nurses, pharmacists, other allied health professionals, and research administrators at 176 NHS hospital organizations across the whole of the UK, supported by staff at the NIHR Clinical Research Network, NHS DigiTrials, Public Health England, Department of Health & Social Care, the Intensive Care National Audit & Research Centre, Public Health Scotland, National Records Service of Scotland, the Secure Anonymised Information Linkage (SAIL) at University of Swansea, and the NHS in England, Scotland, Wales and Northern Ireland. We would especially like to thank the members of the independent Data Monitoring Committee. But above all, we would like to thank the thousands of patients who participated in this study.

